# Influenza vaccine effectiveness against outpatient acute respiratory illness with laboratory-confirmed influenza, United States, 2024–25 season

**DOI:** 10.64898/2026.03.24.26348229

**Authors:** Jessie R. Chung, Ashley M. Price, Stacey L. House, Jamie Mills, Karen J. Wernli, Magali Sanchez, Emily T. Martin, Ivana A. Vaughn, Vel Murugan, Joanna Kramer, Elie A. Saade, Kiran Faryar, Manjusha Gaglani, Chandni Raiyani, Richard K. Zimmerman, Louise H. Taylor, Olivia L. Williams, Emmanuel B. Walter, Juliana DaSilva, Marie K. Kirby, Min Z. Levine, Rebecca Kondor, Emma K. Noble, Kelsey M. Sumner, Sascha R. Ellington, Brendan Flannery, US Flu VE Network Investigators

## Abstract

**Background:** Influenza A(H1N1)pdm09 and A(H3N2) viruses predominated during the 2024–25 U.S. influenza season. We estimated influenza vaccine effectiveness (VE) in the United States against mild-to-moderate outpatient influenza illness by influenza type and subtype in the 2024–25 season.

**Methods:** We enrolled outpatients aged ≥8 months with acute respiratory illness symptoms including cough in 7 states. Upper respiratory specimens were tested for influenza type/subtype by reverse-transcriptase polymerase chain reaction (RT-PCR). Influenza VE was estimated with a test-negative design comparing odds of testing positive for influenza among vaccinated versus unvaccinated participants controlling for age, study site, underlying health status, and month of illness onset. We also estimated VE of current season vaccination among adults stratified by prior season vaccination status.

**Results:** Among 6,793 enrolled patients, 2,016 (30%) tested positive for influenza including 961 A(H3N2), 770 A(H1N1)pdm09, and 183 B/Victoria. Overall vaccine effectiveness against any influenza illness was 33% (95% Confidence Interval [CI]: 24 to 41): 27% (95% CI: 14 to 39) against influenza A(H3N2), 37% (95% CI: 24 to 48) against A(H1N1)pdm09, and 40% (95% CI: 12 to 59) against B/Victoria. VE did not differ based on whether or not participants had received influenza vaccine the previous season.

**Conclusions:** Influenza vaccination during the 2024–25 season protected against circulating influenza viruses, reducing the risk of outpatient medically attended influenza overall by approximately one-third among people who were vaccinated.

**Key Points:** Influenza vaccine reduced the risk of outpatient acute respiratory illness due to laboratory-confirmed influenza during the 2024–25 season by a third.

## Introduction

Influenza causes annual epidemics of acute respiratory illness. During the 2024–25 season, influenza caused an estimated 43 million – 76 million illness episodes, 19 million – 34 million medical visits, 560,000 – 1.2 million hospitalizations and 38,000 – 110,000 influenza-associated deaths in the United States^1^. The 2024–25 influenza season was the first since 2017–18 to be classified as high severity for all age groups^2,3^. For prevention of influenza and complications of influenza virus infection, annual influenza vaccination has been recommended in the United States for all persons aged ≥6 months who do not have contraindications since 2010^4^. Effectiveness of influenza vaccination is monitored to assess protection against circulating influenza viruses and inform annual influenza vaccine updates. Influenza vaccine effectiveness (VE) against medically attended outpatient influenza in the U.S. has ranged from 19% to 60% since 2009^5^. Since 2013, U.S. Centers for Disease Control and Prevention (CDC) has routinely published interim and end-of-season estimates of influenza VE. Annual estimates of the burden averted from influenza vaccination were introduced in 2010. From 2010–11 to 2023–24, influenza vaccination is estimated to have prevented 35 million influenza-associated healthcare visits, 1 million hospitalizations, and 85,000 deaths in the United States ^6^.

In February 2025, interim estimates from four U.S. VE platforms showed significant protection against outpatient and inpatient influenza illness^7^. Here we report updated VE for the 2024–25 season against outpatient medically attended influenza overall by influenza type and A subtype, age group, time since vaccination, and prior season vaccination status.

## Methods

### Study population

The US Flu VE Network has been described previously ^8^. Briefly, this public health surveillance network enrolls participants aged 8 months or older with a mild to moderate severity acute respiratory illness (ARI) ≤7 days duration including new or worsening cough. Patients were enrolled at participating outpatient healthcare facilities, urgent care and emergency departments at sites in Arizona, Michigan, Missouri, Ohio, Pennsylvania, Texas, and Washington. Duke University served as the data coordinating institution for the network. Enrollment ramped up or down at each site based on local evidence of increasing or decreasing influenza activity, respectively (**Supplemental Table 1**). Trained staff interviewed participants or their parent/guardian to obtain demographic and clinical information and inquired about receipt of current season influenza vaccination. Underlying chronic health conditions were defined as either self-report or electronic medical record documentation within the previous year of medical encounters with International Classification of Diseases (ICD-10) codes for high-risk conditions associated with influenza-related complications^9^. The study protocol was reviewed by CDC, determined to be public health surveillance, and conducted consistent with applicable federal law and CDC policy (45 CFR 46.102(l)(2)). Participants or their parent/guardian provided written or oral consent.

### Influenza vaccination

Northern Hemisphere 2024–25 influenza vaccines were trivalent and included two influenza A antigens and one influenza B antigen: A(H3N2) clade 2a.3a.1 subclade J (egg-based A/Thailand/8/2022 or cell-based A/Massachusetts/18/2022), A(H1N1)pdm09 clade 5A.2a.1 subclade C.1.1 (egg-based A/Victoria/4897/2022 or cell-based A/Wisconsin/67/2022) and B/Victoria clade V1A.3a.2 subclade C (egg- or cell-based B/Austria/1359417/2021)^4^. The recommended A(H3N2) vaccine viruses were updated in February 2024; A(H1N1)pdm09 and B/Victoria vaccine viruses were the same as 2023–24 (**Supplemental Table 2**). Sources of influenza vaccine documentation included electronic health records (EHR), state immunization information systems, and vaccine provider information. For patients or parent/guardian reporting children’s vaccination ≥14 days before illness onset, study staff determined plausibility based on reported date and location of vaccination if self-reported vaccination was not captured in electronic immunization records; patients with one or more documented doses of any licensed influenza vaccine product after July 1, 2024 or plausible self-report of influenza vaccination without documentation were considered vaccinated. Influenza vaccine type and prior season (i.e., 2023–24) influenza vaccination status were obtained from electronic immunization records only.

### Laboratory methods

At enrollment, study staff collected nasal and oropharyngeal swabs (nasal only for children younger than 2 years of age) for respiratory virus testing. Respiratory specimens were tested for influenza (including type and influenza A subtype) and SARS-CoV-2 viruses by reverse-transcriptase polymerase chain reaction (RT-PCR) at site laboratories. Case-patients were classified based on study or clinical influenza RT-PCR test result; control-patients were those who tested negative for influenza virus and SARS-CoV-2 infection. Influenza-positive specimens with RT-PCR cycle threshold values <31 were genetically characterized using whole genome sequencing (WGS) at CDC with each sample’s hemagglutinin (HA) clade/subclade assigned using NextClade and based on its consensus sequence^10^.

Serum samples were obtained at enrollment from a subset of patients who agreed to venipuncture. Serum was separated and stored at −20°C or colder at study sites until shipment to CDC for serologic analysis. The microneutralization assay^11^ (MN) was used to quantify antibody levels of A(H3N2)-positive participants at the acute timepoint against the predominant circulating A(H3N2) virus in the Flu VE Network during 2024–25 (A/Wisconsin/154/2024, HA subclade J.2). Assays were conducted as previously described^12^ (**Supplemental Methods**). Control sera from uninfected, influenza-negative participants were matched to case-patient sera based on participant age and month of illness onset. Antibody levels are reported as geometric mean titers (GMT) and 95% confidence intervals. GMTs of those vaccinated and unvaccinated in the prior season were compared using a linear regression model controlling for influenza case status, stratified by current season vaccination status. P-values <0.05 were considered statistically significant.

### Statistical analysis

Influenza VE was estimated with a test-negative design comparing odds of testing positive for influenza among vaccinated versus unvaccinated participants^13^. VE is calculated as (1 – aOR) x 100 expressed as a percent, where aOR is the adjusted odds ratio for RT-PCR-confirmed influenza from multivariable logistic regression models. Models were adjusted a priori for study site, participant age (as age group for all-ages estimates and year of age for age group-specific estimates) at enrollment, presence of ≥1 underlying health conditions (either from documentation or participant/parent/guardian report), and month of illness onset. Additional covariates (sex, race and ethnicity, self-rated general health status, and interval between illness onset and enrollment) were considered for inclusion using a threshold for inclusion of ≥5% change in the aOR. Statistical significance was defined as 95% confidence intervals that did not include zero. We estimated VE overall by influenza A and B type, influenza subtype, and age group (8 months – 4 years, 5–17 years, 8 months – 17 years, 18–49 years, 50–64 years, and ≥65 years). Age group-specific estimates were combined when models did not converge. We estimated VE by age group against any influenza illness by time since vaccination using intervals of 14–60 days, 60–120 days, and >120 days since vaccination compared with unvaccinated participants.

Taking a patient-centered approach^14,15^, we evaluated the added benefit of current season vaccination among patients aged ≥18 years who were vaccinated or unvaccinated the preceding season (2023–24). We used an interaction term of current and prior season vaccination to compare VE estimates^16^. We restricted these analyses to four study sites (MI, PA, TX, and WA) where the majority of that season’s influenza vaccine doses were documented in electronic sources.

## Results

### Participant characteristics

From October 6, 2024–May 10, 2025, 9,782 patients with acute respiratory illness were enrolled at time of presentation for ambulatory care. A total of 2,989 (31%) patients did not meet inclusion criteria for primary VE analyses. Most of the excluded patients already had a clinical influenza diagnostic test result before enrollment which could potentially bias the reported clinical and vaccination information (46% of exclusions). Patients enrolled outside periods of influenza circulation (24% of exclusions) and those who tested RT-PCR positive for SARS-CoV-2 infection (17% of exclusions) also were excluded because influenza and COVID-19 vaccination have been highly correlated^17^ (**Supplemental Figure 1**). Among 6,793 patients who met inclusion criteria, 2,016 (30%) tested positive for influenza virus infection (**Table 1**): 1,830 (91%) influenza A (961 [53%] A(H3N2), 770 [42%] A(H1N1)pdm09, 99 [5%] undetermined subtype), and 183 (9%) for influenza B; 3 (<1%) had influenza A(H1N1)pdm09 and A(H3N2) virus coinfections. Weekly number of influenza-positive participants peaked in week 5 of 2025 (**Supplemental Figure 2**). A total of 2,262 (33%) participants were considered vaccinated against influenza for the 2024–25 season. Of these, 323 (14%) had plausible self-reported vaccination without documentation in electronic immunization record.

**Table 1.** Characteristics of participants enrolled in the US Influenza Vaccine Effectiveness Network for the 2024–25 influenza season.

|  |  | RT-PCR Test result status |  |  |  |  | Influenza Vaccination status |  |  |
| --- | --- | --- | --- | --- | --- | --- | --- | --- | --- |
|  | Total | Influenza-positive |  | Influenza-negative |  |  | Vaccinated |  |  |
| Characteristic | No. | No. | Column % | No. | Column % | p-value | No. | Row % | p-value |
| *Overall* | 6793 | 2016 | 30 | 4777 | 70 |  | 2262 | 33 |  |
| *Study site* |  |  |  |  |  | <0.01 |  |  | <0.01 |
| Arizona | 895 | 296 | 15 | 599 | 13 |  | 120 | 13 |  |
| Michigan | 456 | 116 | 6 | 340 | 7 |  | 225 | 49 |  |
| Missouri | 559 | 64 | 3 | 495 | 10 |  | 150 | 26 |  |
| Ohio | 1120 | 352 | 17 | 768 | 16 |  | 225 | 20 |  |
| Pennsylvania | 1514 | 583 | 29 | 931 | 19 |  | 602 | 40 |  |
| Texas | 1542 | 406 | 20 | 1136 | 24 |  | 589 | 38 |  |
| Washington | 707 | 199 | 10 | 508 | 11 |  | 351 | 50 |  |
| *Sex<sup>b</sup>* |  |  |  |  |  | 0.18 |  |  | <0.01 |
| Female | 3952 | 1148 | 57 | 2804 | 59 |  | 1402 | 36 |  |
| Male | 2819 | 861 | 43 | 1958 | 41 |  | 853 | 30 |  |
| *Age group (years)* |  |  |  |  |  | <0.01 |  |  | <0.01 |
| 8 months–4 years | 1108 | 243 | 12 | 865 | 18 |  | 327 | 29 |  |
| 5–17 | 1299 | 448 | 22 | 851 | 18 |  | 295 | 23 |  |
| 18–49 | 2668 | 854 | 42 | 1814 | 38 |  | 732 | 27 |  |
| 50–64 | 886 | 268 | 13 | 618 | 13 |  | 378 | 42 |  |
| ≥65 | 832 | 203 | 10 | 629 | 13 |  | 530 | 64 |  |
| *Race and ethnicity<sup>c</sup>* |  |  |  |  |  |  |  |  |  |
| American Indian or Alaska Native | 86 | 22 | 1 | 64 | 1 | 0.41 | 27 | 32 | 0.71 |
| Asian | 380 | 129 | 6 | 251 | 5 | 0.06 | 164 | 43 | <0.01 |
| Black or African American | 1862 | 504 | 25 | 1358 | 28 | <0.01 | 464 | 25 | <0.01 |
| Hispanic or Latino | 1057 | 304 | 15 | 753 | 16 | 0.49 | 266 | 25 | <0.01 |
| Middle Eastern or North African | 79 | 32 | 2 | 47 | 1 | 0.03 | 20 | 25 | 0.13 |
| Native Hawaiian or Pacific Islander | 39 | 13 | 1 | 26 | 1 | 0.61 | 13 | 33 | 0.99 |
| White | 3768 | 1131 | 56 | 2637 | 55 | 0.47 | 1472 | 39 | <0.01 |
| *Self-rated general health status<sup>d</sup>* |  |  |  |  |  | <0.01 |  |  | <0.01 |
| Excellent | 1777 | 590 | 29 | 1187 | 25 |  | 502 | 28 |  |
| Very good | 2290 | 709 | 35 | 1581 | 33 |  | 841 | 37 |  |
| Good | 1931 | 552 | 27 | 1379 | 29 |  | 660 | 34 |  |
| Fair/Poor | 772 | 160 | 8 | 612 | 13 |  | 253 | 32 |  |
| *Underlying chronic health conditions<sup>e</sup>* |  |  |  |  |  | <0.01 |  |  | <0.01 |
| None | 3390 | 1105 | 55 | 2285 | 48 |  | 858 | 25 |  |
| ≥1 | 3403 | 911 | 45 | 2492 | 52 |  | 1404 | 41 |  |
| *Illness onset to enrollment (days)* |  |  |  |  |  | <0.01 |  |  | <0.01 |
| <3 | 2838 | 983 | 49 | 1855 | 39 |  | 911 | 32 |  |
| 3–4 | 2307 | 691 | 34 | 1616 | 34 |  | 745 | 32 |  |
| 5–7 | 1648 | 342 | 17 | 1306 | 27 |  | 606 | 37 |  |
| *Received 2023–24 influenza vaccine* | 1935 | 495 | 25 | 1440 | 30 | <0.01 | 1318 | 68 | <0.01 |

<sup>b</sup> Sex was missing for 22 participants.
<sup>c</sup> Participants self-identified all options that applied and could have chosen more than one. 42 participants did not select any of the answer choices.
<sup>d</sup> Self-rated general health status was missing for 23 participants.
<sup>e</sup> Underlying chronic health conditions included chronic cardiac diseases and circulatory diseases, chronic pulmonary diseases, diabetes mellitus, chronic renal disease, hemoglobinopathies, immunosuppressive disorders, malignancy, metabolic diseases, liver diseases, neurological/musculoskeletal conditions, cerebrovascular disease, morbid obesity, and endocrine disorders.

Influenza-positive case patients were more likely to report better general health and sought care earlier compared to test-negative control patients; case patients were less likely to have an underlying health condition than controls (**Table 1**). The proportion of participants vaccinated differed by study site, sex, age group, race and ethnicity, general health status, presence of an underlying health condition, and number of days between illness onset to enrollment (**Table 1**). Most participants vaccinated in 2024–25 (1,318, 68%) were also vaccinated in the 2023–24 season. Among participants aged <65 years, the type of influenza vaccine received was documented in electronic immunization records for 1,295 (75%) of 1,732 vaccinated patients: 815 (63%) received egg-based inactivated influenza vaccine (807 [99%] standard dose [IIV3-SD], 5 [1%] live-attenuated influenza vaccine, 2 [<1%] high-dose [IIV3-HD], and1 [<1%] adjuvanted [aIIV3]), and 480 (37%) received non-egg-based vaccines (424 [88%] cell culture-based IIV3 [ccIIV3] and 56 [12%] recombinant influenza vaccine [RIV3]). Among adults aged ≥65 years at enrollment, influenza vaccine type was documented for 464 (88%) of 530 vaccinated participants: 439 (95%) received a preferentially recommended vaccine^4^ (235 [54%] IIV3-HD, 179 [41%] aIIV3, and 25 [6%] RIV3).

### Genetic characterization

Of 1,478 influenza-positive respiratory specimens that met criteria for WGS, 1,380 (93%) viruses were successfully sequenced: 692 A(H3N2), 571 A(H1N1)pdm09 and 117 B/Victoria (**Supplemental Table 3**). A(H1N1)pdm09 viruses belonged to two HA clades: 406 (71%) to 5a.2a.1 and 165 (29%) to 5a.2a. All A(H3N2) viruses belonged to the 2a.3a.1 HA clade; most (479, 69%) belonged to subclade J.2, and smaller proportions belonged to J.2.2 (111, 16%), J.2.5 (77, 11%), J.2.1 (6, 1%), J.2.4 (8, 1%), or J.1.1 (1, <1%). All influenza B/Victoria viruses belonged 3a.2 HA clade C: 43% subclade C.3.1, 27% C.5.1, 11% C.5.6, 9% C.5.7, 4% C.5.6.1, 3% C.5, and 2% C.3.2.

### Overall influenza vaccine effectiveness

For the period from October 2024 through May 2025, the overall VE against medically attended outpatient influenza A and B viruses was 33% (95% confidence interval [CI]: 24 to 41); VE was not statistically significant among patients aged ≥65 years (VE: 4%, 95%CI: −42 to 35) (**Table 2**). VE was 27% (95%CI: 14 to 39) against A(H3N2), 37% (95%CI: 24 to 48) against influenza A(H1N1)pdm09, and 40% (95%CI: 12 to 59) against influenza B/Victoria. VE against influenza A(H3N2) and A(H1N1)pdm09 was highest among children aged 8 months to 4 years (41% and 73%, respectively).

**Table 2.** Influenza vaccine effectiveness against outpatient influenza-associated illness visits among patients aged ≥8 months enrolled at US Influenza Vaccine Effectiveness Network sites, October 2024 through May 2025.

|  | Influenza Positive (Cases) |  | Influenza Negative (Controls) |  | Vaccine Effectiveness<sup>a</sup> |  |
| --- | --- | --- | --- | --- | --- | --- |
|  | No. vaccinated/Total | % Vaccinated | No. vaccinated/Total | % Vaccinated | VE % | (95% CI) |
| **All influenza viruses** |  |  |  |  |  |  |
| All ages ≥8 months | 575/2016 | 29 | 1687/4777 | 35 | 33 | (24 to 41) |
| 8 months–17 years | 146/691 | 21 | 476/1716 | 28 | 41 | (25 to 53) |
| 8 months–4 years | 51/243 | 21 | 276/865 | 32 | 50 | (29 to 66) |
| 5–17 | 95/448 | 21 | 200/851 | 24 | 26 | (-1 to 45) |
| 18–49 | 205/854 | 24 | 527/1814 | 29 | 31 | (15 to 44) |
| 50–64 | 99/268 | 37 | 279/618 | 45 | 37 | (12 to 55) |
| ≥65 | 125/203 | 62 | 405/629 | 64 | 4 | (-42 to 35) |
| **Influenza A(H3N2)** |  |  |  |  |  |  |
| All ages ≥8 months | 269/964 | 28 | 1687/4777 | 35 | 27 | (14 to 39) |
| 8 months–17 years | 69/333 | 21 | 476/1716 | 28 | 42 | (21 to 57) |
| 8 months–4 years | 31/131 | 24 | 276/865 | 32 | 41 | (7 to 63) |
| 5–17 | 38/202 | 19 | 200/851 | 24 | 36 | (1 to 58) |
| 18–49 | 101/437 | 23 | 527/1814 | 29 | 24 | (1 to 42) |
| 50–64 | 40/104 | 38 | 279/618 | 45 | 33 | (-9 to 58) |
| ≥65 | 59/90 | 66 | 405/629 | 64 | -29 | (-123 to 25) |
| **Influenza A(H1N1)pdm09** |  |  |  |  |  |  |
| All ages ≥8 months | 237/773 | 31 | 1687/4777 | 35 | 37 | (24 to 48) |
| 8 months–17 years | 46/238 | 19 | 476/1716 | 28 | 54 | (33 to 68) |
| 8 months–4 years | 12/83 | 14 | 276/865 | 32 | 73 | (47 to 86) |
| 5–17 | 34/155 | 22 | 200/851 | 24 | 30 | (-12 to 56) |
| 18–49 | 81/297 | 27 | 527/1814 | 29 | 25 | (-2 to 45) |
| 50–64 | 50/136 | 37 | 279/618 | 45 | 41 | (9 to 32) |
| ≥65 | 60/102 | 59 | 405/629 | 64 | 18 | (-35 to 50) |
| **Influenza B/Victoria** |  |  |  |  |  |  |
| All ages ≥8 months | 44/183 | 24 | 1687/4777 | 35 | 40 | (12 to 59) |
| 8 months–17 years | 26/96 | 27 | 476/1716 | 28 | 25 | (-27 to 55) |
| ≥18 years | 18/87 | 21 | 1211/3061 | 40 | 63 | (34 to 79) |

Abbreviations: CI, confidence interval; VE, vaccine effectiveness
<sup>a</sup> Models adjusted for study site, age, presence of ≥1 underlying health condition, and month of illness onset. 95% confidence intervals that exclude 0% are considered statistically significant.
<sup>b</sup> Total influenza cases include 99 influenza A-positives of undetermined subtype and 3 influenza A(H1N1)pdm09 and A(H3N2) coinfections. Coinfections are included as cases of both detected viruses in the table.

### Effects of time since influenza vaccination

The median time between influenza vaccination and illness onset was 108 days (interquartile range [IQR] 65–146 days). Overall and for each age group, VE point estimates were highest among participants vaccinated 14–59 days prior to illness onset compared to those with longer intervals although confidence intervals overlapped (**Supplemental Table 4**).

### Effects of prior season influenza vaccination

Among 2,637 participants from the four sites included in the analysis, 817 (31%) individuals were vaccinated against influenza in both the current (2024–25) and prior (2023–24) seasons, 442 (17%) were vaccinated in the current season only, 224 (8%) were vaccinated in the prior season only, and 1,154 (44%) were unvaccinated in both seasons (**Table 3**). Participants unvaccinated in both seasons were more likely to test positive compared to other groups. Among participants who were not vaccinated in the prior season, VE of current season vaccination against any influenza was 33% (95% CI: 13 to 49) compared with 17% (95%CI: −20 to 43) among participants who were vaccinated in both seasons (p-value for interaction = 0.26). VE against A(H3N2) or A(H1N1)pdm09 did not differ by prior season vaccination status (p-values for interaction 0.81 and 0.32, respectively). Sample sizes were insufficient to estimate VE against influenza B/Victoria by prior vaccination status.

**Table 3.**
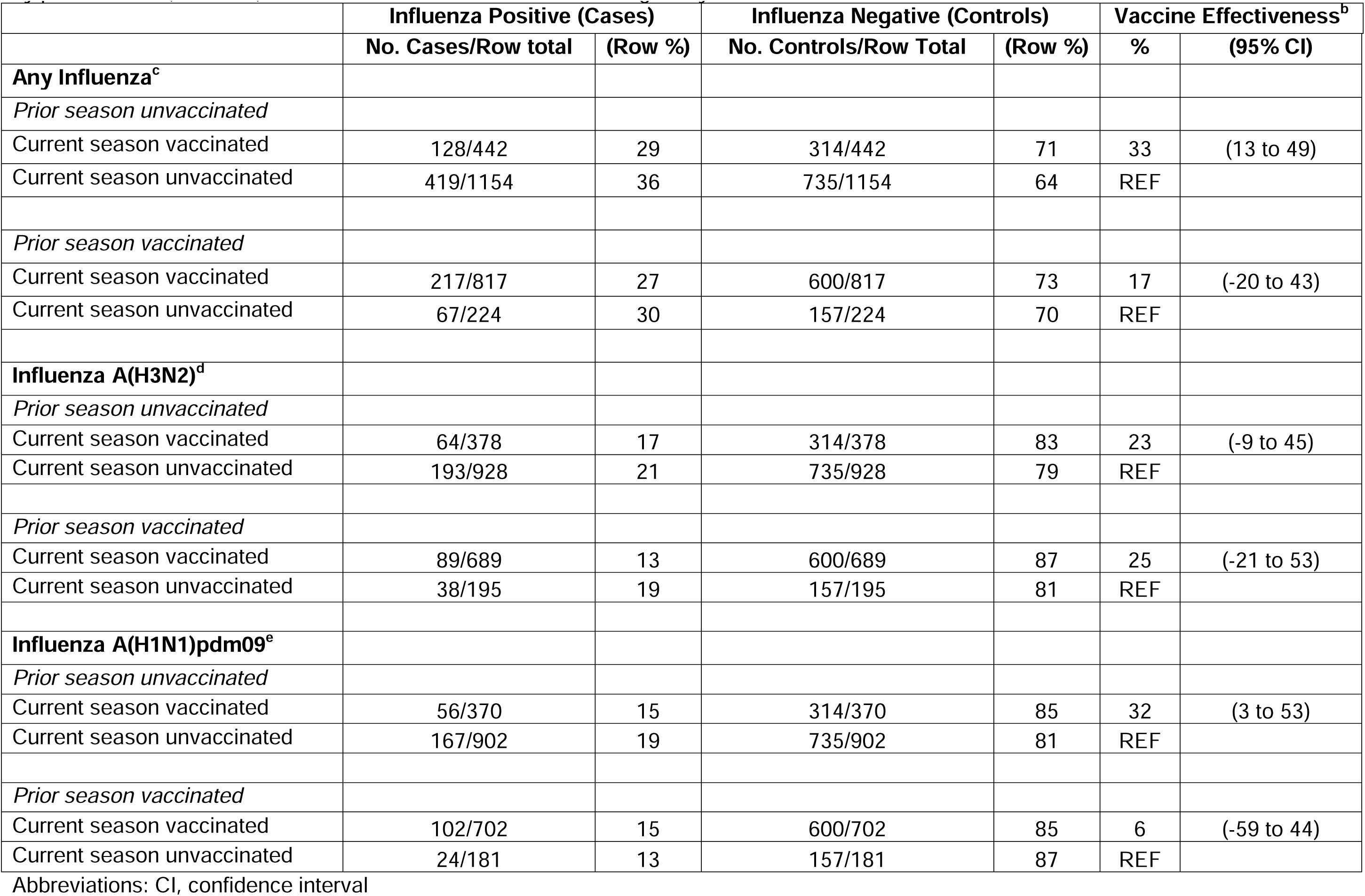
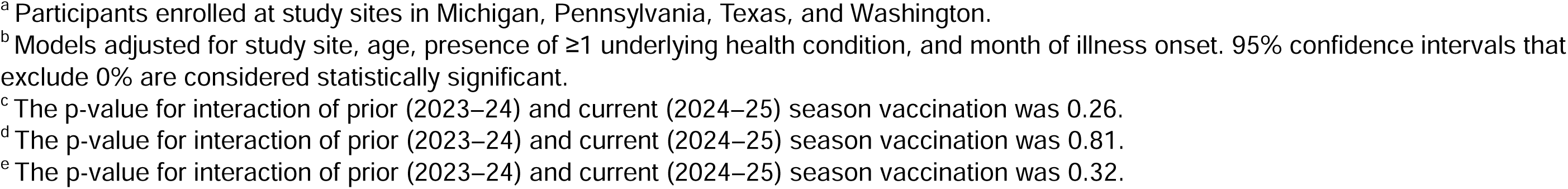
Influenza vaccine effectiveness against outpatient influenza-associated illness visits among participants^a^ aged ≥18 years stratified by prior season (2023–24) vaccination status, October 2024 through May 2025.

|  | Influenza Positive (Cases) |  | Influenza Negative (Controls) |  | Vaccine Effectiveness<sup>b</sup> |  |
| --- | --- | --- | --- | --- | --- | --- |
|  | No. Cases/Row total | (Row %) | No. Controls/Row Total | (Row %) | % | (95% CI) |
| **Any Influenza<sup>c</sup>** |  |  |  |  |  |  |
| *Prior season unvaccinated* |  |  |  |  |  |  |
| Current season vaccinated | 128/442 | 29 | 314/442 | 71 | 33 | (13 to 49) |
| Current season unvaccinated | 419/1154 | 36 | 735/1154 | 64 | REF |  |
| *Prior season vaccinated* |  |  |  |  |  |  |
| Current season vaccinated | 217/817 | 27 | 600/817 | 73 | 17 | (-20 to 43) |
| Current season unvaccinated | 67/224 | 30 | 157/224 | 70 | REF |  |
| **Influenza A(H3N2)<sup>d</sup>** |  |  |  |  |  |  |
| *Prior season unvaccinated* |  |  |  |  |  |  |
| Current season vaccinated | 64/378 | 17 | 314/378 | 83 | 23 | (-9 to 45) |
| Current season unvaccinated | 193/928 | 21 | 735/928 | 79 | REF |  |
| *Prior season vaccinated* |  |  |  |  |  |  |
| Current season vaccinated | 89/689 | 13 | 600/689 | 87 | 25 | (-21 to 53) |
| Current season unvaccinated | 38/195 | 19 | 157/195 | 81 | REF |  |
| **Influenza A(H1N1)pdm09<sup>e</sup>** |  |  |  |  |  |  |
| *Prior season unvaccinated* |  |  |  |  |  |  |
| Current season vaccinated | 56/370 | 15 | 314/370 | 85 | 32 | (3 to 53) |
| Current season unvaccinated | 167/902 | 19 | 735/902 | 81 | REF |  |
| *Prior season vaccinated* |  |  |  |  |  |  |
| Current season vaccinated | 102/702 | 15 | 600/702 | 85 | 6 | (-59 to 44) |
| Current season unvaccinated | 24/181 | 13 | 157/181 | 87 | REF |  |

Abbreviations: CI, confidence interval
<sup>a</sup> Participants enrolled at study sites in Michigan, Pennsylvania, Texas, and Washington.
<sup>b</sup> Models adjusted for study site, age, presence of $\geq 1$ underlying health condition, and month of illness onset. 95% confidence intervals that exclude 0% are considered statistically significant.
<sup>c</sup> The p-value for interaction of prior (2023–24) and current (2024–25) season vaccination was 0.26.
<sup>d</sup> The p-value for interaction of prior (2023–24) and current (2024–25) season vaccination was 0.81.
<sup>e</sup> The p-value for interaction of prior (2023–24) and current (2024–25) season vaccination was 0.32.

### Serum antibody levels to influenza A(H3N2)

Sera from 232 adults (102 A(H3N2)-positive participants and 130 influenza-negative participants) were tested by the microneutralization assay (**Supplemental Table 5**). Acute-phase antibody titers against the predominant circulating A(H3N2) virus from HA subclade J.2, A/Wisconsin/154/2024, were higher among test-negative control participants (GMT: 19, 95%CI: 15 to 24) than A(H3N2)-positive cases (GMT: 10, 95%CI: 8 to 12). Among test-negative controls, participants who were vaccinated in the current season had higher antibody levels (GMT: 44, 95%CI: 28 to 68) compared to unvaccinated participants (GMT: 14, 95%CI: 11 to 18). Stratifying by current season vaccination status and controlling for influenza case status, GMT was not associated with prior season vaccination status (p=0.40 for those vaccinated in the current season and p=0.74 for those who did not receive the current season vaccination).

### Sensitivity analyses

VE against any influenza using self- or parent/guardian-reported influenza vaccination only was 23% (95%CI: 11 to 33); VE using EHR-documented data only was 37% (95%CI: 28 to 45) (**Supplemental Table 6**). Inclusion of participants who could have known their clinical influenza test result at the time of enrollment reduced VE to 28% (95%CI: 19 to 35). All other sensitivity analyses varying inclusion criteria for analysis were all within 6 percentage points of the overall VE reported in the primary analysis.

## Discussion

During the 2024–25 influenza season in the U.S., with co-circulation of influenza A(H3N2) and A(H1N1)pdm09 viruses, influenza vaccination reduced the risk of outpatient medically attended influenza by approximately one-third. Influenza vaccination was associated with lower odds of A(H3N2)-, A(H1N1)pdm09-, and B/Victoria-associated illness with variable levels of protection observed by participant age category. VE may have been affected by genetic and antigenic differences between influenza vaccine components and circulating influenza viruses. Statistically significant protection against medically attended influenza was observed up to 4 months after vaccination with 2024–25 vaccine. VE did not differ by prior season influenza vaccination status, which was supported by similar serum antibody levels against the predominant circulating A(H3N2) virus in participants who were and were not vaccinated against influenza in the prior season. Genetic characterization of influenza viruses and measurement of antibody levels contributed to understanding of protection provided by influenza vaccination in this season.

Seasonal influenza vaccination has been associated with higher protection against A(H1N1)-associated illness compared to protection against A(H3N2). In meta-analyses of VE studies against medically attended outpatient illness, pooled VE against A(H3N2) among all persons ranged from 22–33%, and VE against A(H1N1)pdm09 ranged from 56–61%^18,19^. During 2024–25, influenza vaccination provided protection against A(H1N1)pdm09- and A(H3N2)-associated outpatient visits, with similar estimates of overall VE. Estimated VE against A(H3N2) fell within the range of recent US Flu VE Network estimates during A(H3N2)-predominant seasons. However, VE against A(H1N1)pdm09 was lower than has been seen during seasons when circulating A(H1N1)pdm09 viruses were antigenically similar to vaccine components. During the 2019–20 season, the US Flu VE Network reported decreased protection against an antigenically drifted A(H1N1)pdm09 virus^20^. Observed protection in this Network has been <50% in the subsequent seasons with A(H1N1)pdm09 circulation.

In the 2024–25 season, children in the youngest age group had the highest VE point estimates against influenza A, which is consistent with prior analyses and other studies^21,22^. Among adults aged ≥65 years, who are at higher risk of influenza-associated complications, multiple studies demonstrated protection against influenza A in the 2024–25 season^7^. In this analysis, VE against influenza A was not statistically significant. VE against A(H3N2) in this age group has been consistently low; in meta-analyses, pooled VE against A(H3N2) was 11–24% among older adults^18,19^. The A(H3N2) vaccine components were updated for the 2025–26 season^23^ (**Supplemental Table 2**). Estimated VE against A(H1N1)pdm09 was lower among older adults compared to pooled meta-analyses estimates (47–62%)^18,19^ but was overlapping with the estimate from the US Flu VE Network from the 2023–24 season (39%, 95%CI: 2 to 62)^8^. In the United States, uptake of adjuvanted and higher-dose influenza vaccines has increased since these vaccines were preferentially recommended for adults aged ≥65 years. However, during the current season when most adults aged ≥65 years received adjuvanted or higher dose vaccines, VE was comparable to prior seasons, despite evidence of improved immunogenicity and effectiveness in some studies^24–26^. Waning immunity might be a contributing factor because individuals aged ≥65 years may be more likely to seek vaccination early^27^. In our study, fewer adults aged ≥65 years were vaccinated within 60 days of illness onset compared to adults aged 18–64 years. Influenza B detections were more numerous among children than adults, and observed VE was lower among children than has been found in prior seasons. Our study included a larger proportion of viruses belonging to the emerging B/Victoria HA subclade C.3.1 (43%) compared to national surveillance data for the season (12.9% of sequenced B/Victoria viruses)^3^, which may have contributed to lower VE against B/Victoria.

We did not detect differences in VE by prior season influenza vaccination status overall or against A(H3N2) or A(H1N1)pdm09 among adults from four study sites with the most complete electronic vaccination records. Higher VE estimates among people vaccinated against influenza only in the current season compared to repeat vaccinees (i.e., those vaccinated in both current and prior seasons) have been reported in several studies^28–30^. In our study, repeat vaccination effects were not statistically significant. Serum antibody titers against the predominantly circulating A(H3N2) virus were consistent with similar protection against laboratory-confirmed influenza gained by current season vaccination regardless of prior season vaccination status. Analyses from the patient perspective over multiple seasons have shown more consistent evidence of the added benefit of current season influenza vaccination even among people vaccinated during the preceding season regardless of antigenic similarities between vaccine components and circulating viruses over two seasons^14,15^.

Findings are subject to several limitations. First, current and prior season influenza vaccination may not have been documented leading to misclassification of vaccinated patients as unvaccinated and underestimating VE. However, documentation was available for 84% of patients with plausible self-report of current season vaccination. Some patients vaccinated against influenza in the prior season may have been inadvertently included in the unvaccinated group in analyses comparing the effect of prior season vaccination. Analyses of prior season vaccine effects were restricted to study sites with high proportions of documented vaccinations. Second, while the test-negative design reduces potential bias due to differences in healthcare seeking, unmeasured confounding related to study enrollment may have affected VE estimates. Findings were robust to changes in inclusion criteria and definitions of exposure status. Patients were systematically tested by highly sensitive and specific RT-PCR for influenza and SARS-CoV-2, reducing misclassification of case status. Enrolled patients were not tested systematically for Respiratory Syncytial Virus (RSV) infection. Missing data on RSV infection were unlikely to have affected influenza VE estimates. As RSV vaccine uptake increases among populations in which vaccination is recommended, systematic testing for RSV in the future may be considered to remove potential bias^31,32^. Third, we did not compare full^4^ versus partial influenza vaccination among children aged <9 years. Receipt of the recommended number of doses has been associated with higher VE among young children compared to partial vaccination^33^. Finally, we did not have serologic testing results for A(H1N1)pdm09- or influenza B-positive cases due to resource constraints.

The results presented in this study demonstrate an overall benefit of receiving the 2024–25 influenza vaccine in the United States; vaccination reduced the risk of outpatient medically attended influenza overall by approximately one-third. In seasons such as 2024–25, when the burden of influenza is high, even modestly effective vaccines can still prevent substantial numbers of influenza-associated medical visits, hospitalizations, and deaths^34,35^.

## Data Availability

All data produced in the present study are available upon reasonable request to the authors.

## Funding and Conflicts of interest

The US Flu VE Network is funded through a US Centers for Disease Control and Prevention Cooperative Agreement (5U01 IP001180-03, 5U01 IP001181-03, 5U01 IP001182-03, 5U01 IP001184-03, 5U01 IP001189-03, 5U01 IP001191-03, 5U01 IP001193-03, and 5U01 IP001194-03). The University of Pittsburgh site was also supported by National Institutes of Health grant UL1TR001857.

SLH has received grants from Seegene Inc., Abbott, Healgen, Roche, CorDx, Hologic, Cepheid, Janssen, Roche, Alfa Scientific, Nuclein, CDx Diagnostics, and Wondfo Biotech. KJW reports research funding from Eli Lilly, unrelated to the topic under study. ETM has received grants from Merck. IAV has received funding through her institution from eMaxHealth, Evidera, PPD, Vantive Health LLC, UCB Biosciences, Boehringer Ingelheim, and Eli Lilly for unrelated research projects. EAS has received grants from Protein Sciences Corporation and consulting fees from Johnson and Johnson. MJG has received grants from CDC-Vanderbilt Medical Center, CDC-Abt Associates and CDC-Westat for respiratory virus epidemiology and vaccine effectiveness studies. RKZ has received grants and consulting from Sanofi Pasteur and Merck. EBW has received research funding from Pfizer, Moderna, Seqirus, Najit Technologies, and Clinetic for the conduct of clinical research studies. He has also received support as an advisor to Vaxcyte, Seqirus, and Pfizer consultant to ILiAD Biotechnologies, and DSMB member for Shionogi and Emmes. All other authors report nothing to disclose.

## Disclaimer

The findings and conclusions in this report are those of the authors and do not necessarily represent the official position of the Centers for Disease Control and Prevention.

## US Flu VE Network Investigators

Tamair Curley, Jeremy Russell, Ben Rapp, and Joshua Jackson (Washington University School of Medicine in St. Louis); Erika Kiniry, Brianna Wickersham, Rachael Doud, and Matthew Nguyen (Kaiser Permanente Washington Health Research Institute); Arnold S. Monto, Donny Hearn III, and Caroline K. Cheng (University of Michigan); Maria Santana-Garces, Muniza Hossain, and Shane Bole (Henry Ford Health); Mehal Patel (Phoenix Children’s Hospital); Lora Nordstrom (Valleywise Health); Matthew Scotch, Bradley Bobbett (Arizona State University); Alexia El Khoury, Joy Abou Farah, (University Hospitals of Cleveland) Tracy Lemonovich, (MetroHealth), Curtis Donskey (Louis Stokes Cleveland Veterans Affairs Medical Center); Spencer Rose, Michael Smith, Leah Odame-Bamfo, Kempapura Murthy, Mufaddal Mamawala, Amanda McKillop, Nicole Calhoun, Britan Fairall (Baylor Scott & White Health); Mary Patricia Nowalk, GK Balasubramani, Tracey Conti, David A. Figucia, and Alexandra Weissman (University of Pittsburgh); Natalie A. B. Bontrager, Darren J. Morrow, Wes Rountree, Christopher A. Todd, and Cameron R. Wolfe (Duke University); Lisa Keong, Sydney R. Sheffield, Julia C. Frederick, Malania M. Wilson, Ewelina Lyszkowicz, Yujin Jung, Crystal Holiday, Stacie Jefferson, and Philip Shirk (US Centers for Disease Control and Prevention)

## Acknowledgements

We thank the participants of the US Flu VE Network. The Arizona State University team acknowledges the contributions of Kelly Conard, Ching-Wen Hou, Lucy Sublasky-Rodriguez, LaRinda Holland, Izamar Garcia, Giavanna Caruth, Alexis Thomas, Gabrielle Hernandez Barrera, Jesus Estrada, Karen M. Yeager, Raquel Salgado, Martine Desulme and Bryan Remuto; The Baylor Scott & White Health team acknowledges the contributions of, Ashley Graves, Martha Zayed, Marcus Volz, Tora Adams, Jenica Aldas, Vanessa Crumpton, Erica Hayes, Paula Harkins, Victoria Harkins-Walston, Maria Lopez, Arnesa Pepino, Seretha Wallace, Sharla Russell, Briget Da Graca, Jason Ramm, Muralidhar Jatla, Madhava Beeram, Tresa McNeal, Keith Stone, Arundhati Rao, Manohar Mutnal, Dedra Preece, Jason Ettlinger, Courtney Shaver, Monica Bennett, Elisa Priest, Jennifer Thomas, Javed Butler and Alejandro Arroliga. Kaiser Permanente Washington further acknowledges the contributions of Stephen Alexanian, Katherine Chen, Joe Choe, Rachael Doud, Victor Garcia, Rachael Lucero, Kathryn Moser, Paul Thottingal, and Brian Williamson. The University of Michigan group acknowledges the contributions of Miren Oraa, Jacoby Jennings, Emileigh Johnson, Lara J. Thomas, Anne Kaniclides, Dolapo Raji, Leo Sullivan, Ana Montaner, Rachel Truscon, Mona Albaity, Shifa Lakhani, Dirk (Tyler) Thornhill, and James Kwag. The University of Pittsburgh group acknowledges the contributions of Michael Susick, Helen D’Agostino, Robert Hickey, Joe Suyama, Laura Herrmann, Minnie Quebral, Gabriella Alicea, Peyton Sims, and Monika Johnson. The Washington University School of Medicine team acknowledges the contributions of the staff of the Emergency Care Research Core including Cindy Charles, Anna Davidson, Serena House, Samuel Kurtz, Heena Khan, Taylor Kristicevich, Michael Lehmkuhl, Rianna Robinson, Ian Schamburg, Venice Vega, Sarah Welsh, Wykita Willis-Lockhart, and Jessica Zvanut.

---

**Supplemental Table 1.** Enrollment dates included by site.

| **State** | **Enrollment Institutions** | **Enrollment start date** | **Enrollment end date** |
| --- | --- | --- | --- |
| Arizona | Arizona State University,<br>Valleywise Health,<br>Phoenix Children's Hospital | October 13, 2024 | April 28, 2025 |
| Michigan | University of Michigan,<br>Henry Ford Health | December 9, 2024 | May 10, 2025 |
| Missouri | Barnes Jewish Memorial Hospital,<br>St. Louis Children's Hospital | October 7, 2024 | April 23, 2025 |
| Ohio | University Hospitals at Cleveland,<br>Cleveland Veterans' Health Administration | October 31, 2024 | March 26, 2025 |
| Pennsylvania | University of Pittsburgh Medical Center,<br>Children's Hospital of Pittsburgh | October 30, 2024 | May 3, 2025 |
| Texas | Baylor Scott & White Health | October 6, 2024 | May 10, 2025 |
| Washington | Kaiser Permanente Washington | October 20, 2024 | May 10, 2025 |

**Supplemental Table 2.** US influenza vaccine strains, 2023–24 through 2025–26 seasons.

|  | 2023–24<sup>1</sup> | 2024–25<sup>2</sup> | 2025–26<sup>3</sup> |
| --- | --- | --- | --- |
| **Egg-based vaccines** |  |  |  |
| A(H3N2) | A/Darwin/9/2021 | A/Thailand/8/2022 | A/Croatia/10136RV/2023 |
| A(H1N1)pdm09 | A/Victoria/4897/2022 | A/Victoria/4897/2022 | A/Victoria/4897/2022 |
| B/Victoria | B/Austria/1359417/2021 | B/Austria/1359417/2021 | B/Austria/1359417/2021 |
| B/Yamagata | B/Phuket/3073/2013 | Not applicable | Not applicable |
| **Cell culture-based and recombinant vaccines** |  |  |  |
| A(H3N2) | A/Darwin/6/2021 | A/Massachusetts/18/2022 | A/District of Columbia/27/2023 |
| A(H1N1)pdm09 | A/Wisconsin/67/2022 | A/Wisconsin/67/2022 | A/Wisconsin/67/2022 |
| B/Victoria | B/Austria/1359417/2021 | B/Austria/1359417/2021 | B/Austria/1359417/2021 |
| B/Yamagata | B/Phuket/3073/2013 | Not applicable | Not applicable |

<sup>1</sup> https://www.who.int/news/item/24-02-2023-recommendations-announced-for-influenza-vaccine-composition-for-the-2023-2024-northern-hemisphere-influenza-season
<sup>2</sup> https://www.who.int/news/item/23-02-2024-recommendations-announced-for-influenza-vaccine-composition-for-the-2024-2025-northern-hemisphere-influenza-season
<sup>3</sup> https://www.who.int/publications/m/item/recommended-composition-of-influenza-virus-vaccines-for-use-in-the-2025-2026-nh-influenza-season

**Supplemental Figure 1.**
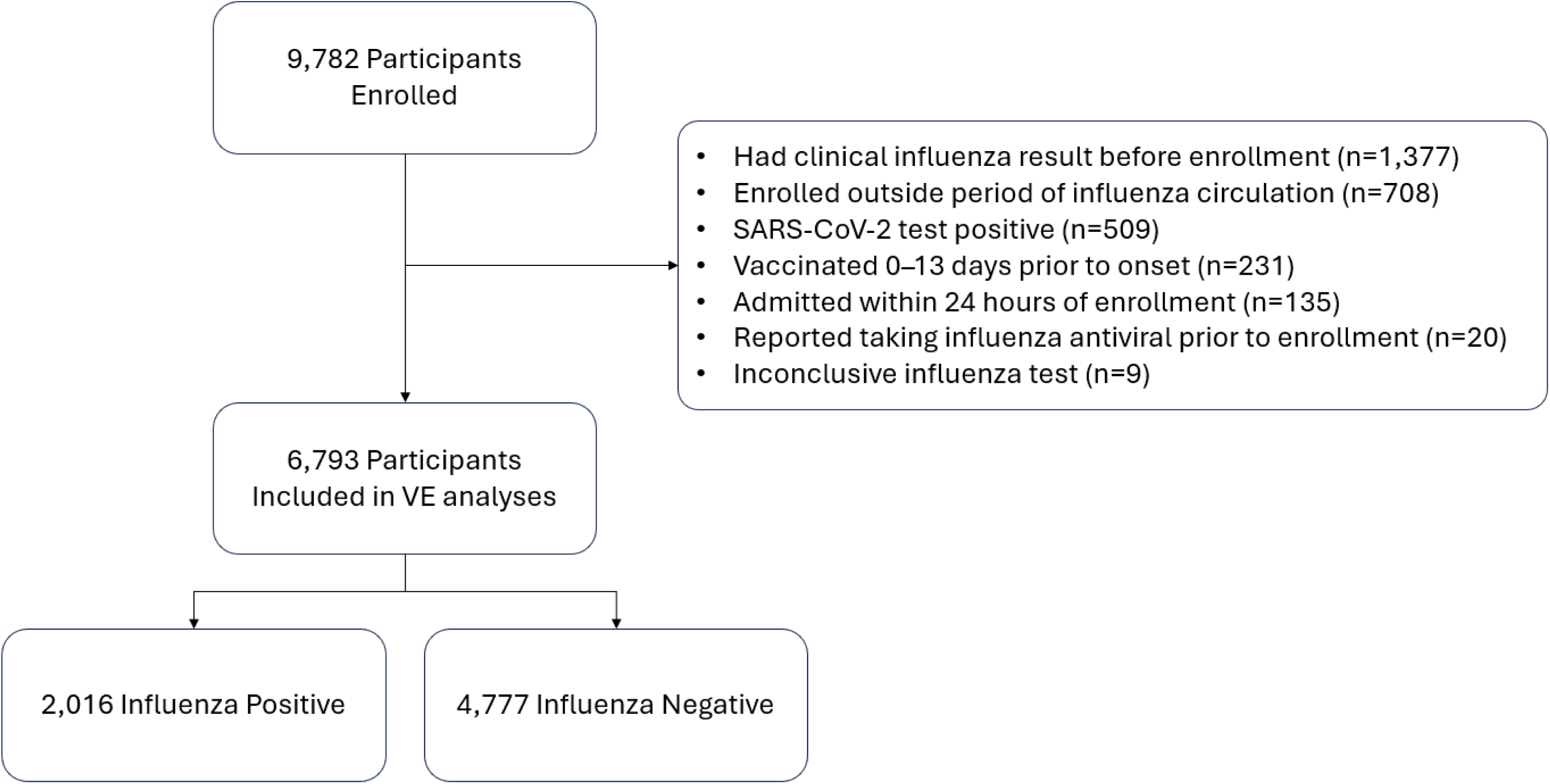
Exclusion criteria figure

**Supplemental Figure 2.**
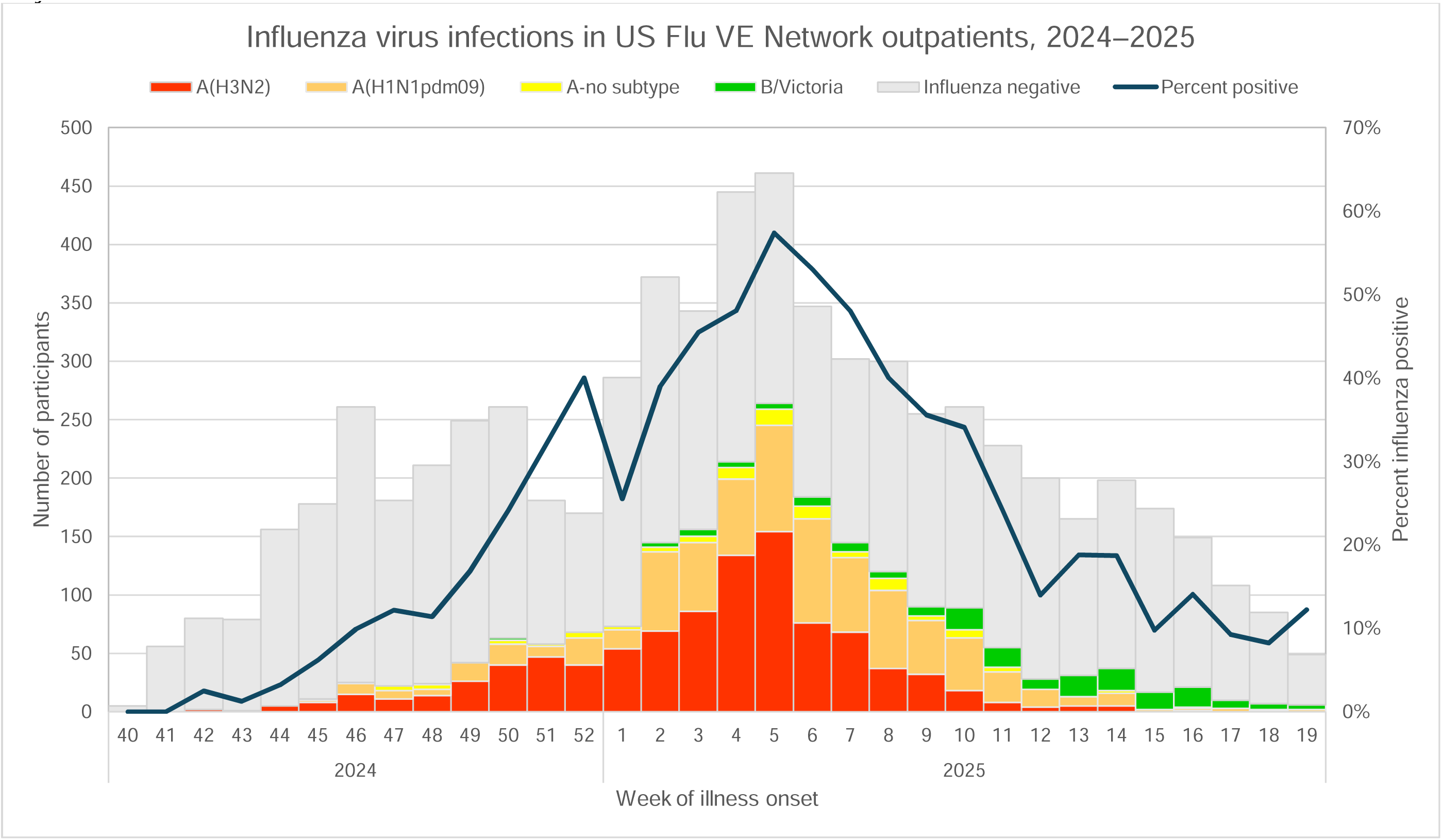
Influenza-positive cases by type/subtype and percent influenza positive by week of enrollment, October 2024 – May 2025.

**Supplemental Table 3.** Whole-genome sequencing results, US Flu VE Network, October 2024 – May 2025.

| Influenza Virus Subtype or Lineage | Genetic Characterization |  |  |  |  |
| --- | --- | --- | --- | --- | --- |
|  | Total No. of Subtype/Lineage Tested | HA Clade | Number (% of subtype/lineage tested) | HA Subclade | Number (% of clade tested) |
| **A(H1N1)pdm09** | 571 | 5a.2a | 165 (29%) | C.1.9 | 53 (32%) |
|  |  |  |  | C.1.9.1 | 14 (8%) |
|  |  |  |  | C.1.9.2 | 5 (3%) |
|  |  |  |  | C.1.9.3 | 93 (56%) |
|  |  | 5a.2a.1 | 406 (71%) | D | 8 (2%) |
|  |  |  |  | D.1 | 1 (0.2%) |
|  |  |  |  | D.3 | 359 (88%) |
|  |  |  |  | D.5 | 39 (10%) |
| **A(H3N2)** | 692 | 2a.3a.1 | 692 (100%) | J.1.1 | 1 (0.1%) |
|  |  |  |  | J.2 | 479 (69%) |
|  |  |  |  | J.2.1 | 6 (1%) |
|  |  |  |  | J.2.2 | 111 (16%) |
|  |  |  |  | J.2.3 | 10 (1%) |
|  |  |  |  | J.2.4 | 8 (1%) |
|  |  |  |  | J.2.5 | 77 (11%) |
| **B/Victoria** | 117 | 3a.2 | 117 (100%) | C.3.1 | 50 (43%) |
|  |  |  |  | C.3.2 | 2 (2%) |
|  |  |  |  | C.5 | 4 (3%) |
|  |  |  |  | C.5.1 | 32 (27%) |
|  |  |  |  | C.5.6 | 13 (11%) |
|  |  |  |  | C.5.6.1 | 5 (4%) |
|  |  |  |  | C.5.7 | 11 (9%) |

**Supplemental Table 4.** Influenza vaccine effectiveness against any outpatient influenza-associated illness visits among patients aged ≥8 months enrolled at US Influenza Vaccine Effectiveness Network sites, October 2024 through May 2025 by time since vaccination.

| Age group | Influenza Positive (Cases) |  | Influenza Negative (Controls) |  | Vaccine Effectiveness<sup>a</sup> |  |
| --- | --- | --- | --- | --- | --- | --- |
|  | Total | No. Vaccinated (%) | Total | No. Vaccinated (%) | VE % | (95% CI) |
| **All ages ≥8 months** |  |  |  |  |  |  |
| 14–59 | 61/1466 | 4 | 358/3403 | 11 | 56 | (40 to 67) |
| 60–120 | 200/1605 | 12 | 524/3569 | 15 | 41 | (28 to 51) |
| >120 | 216/1621 | 13 | 580/3625 | 16 | 16 | (-3 to 31) |
| **8 months –17 years** |  |  |  |  |  |  |
| 14–59 | 26/571 | 5 | 128/1367 | 9 | 55 | (28 to 71) |
| 60–120 | 58/603 | 10 | 172/1411 | 12 | 48 | (28 to 63) |
| >120 | 56/601 | 9 | 160/1399 | 11 | 19 | (-16 to 44) |
| **18–64 years** |  |  |  |  |  |  |
| 14–59 | 28/811 | 3 | 155/1737 | 9 | 56 | (32 to 72) |
| 60–120 | 99/882 | 11 | 233/1815 | 13 | 42 | (24 to 56) |
| >120 | 95/878 | 11 | 249/1831 | 14 | 27 | (2 to 45) |
| **≥65 years** |  |  |  |  |  |  |
| 14–59 | 7/84 | 8 | 75/299 | 25 | 47 | (-32 to 79) |
| 60–120 | 43/120 | 36 | 119/343 | 35 | 5 | (-59 to 43) |
| >120 | 65/142 | 46 | 171/395 | 43 | -35 | (-131 to 21) |

Abbreviations: CI, confidence interval; VE, vaccine effectiveness
<sup>a</sup> Models adjusted for study site, age, presence of ≥1 underlying health condition, and month of illness onset. 95% confidence intervals that exclude 0% are considered statistically significant.

**Supplemental Table 5.**
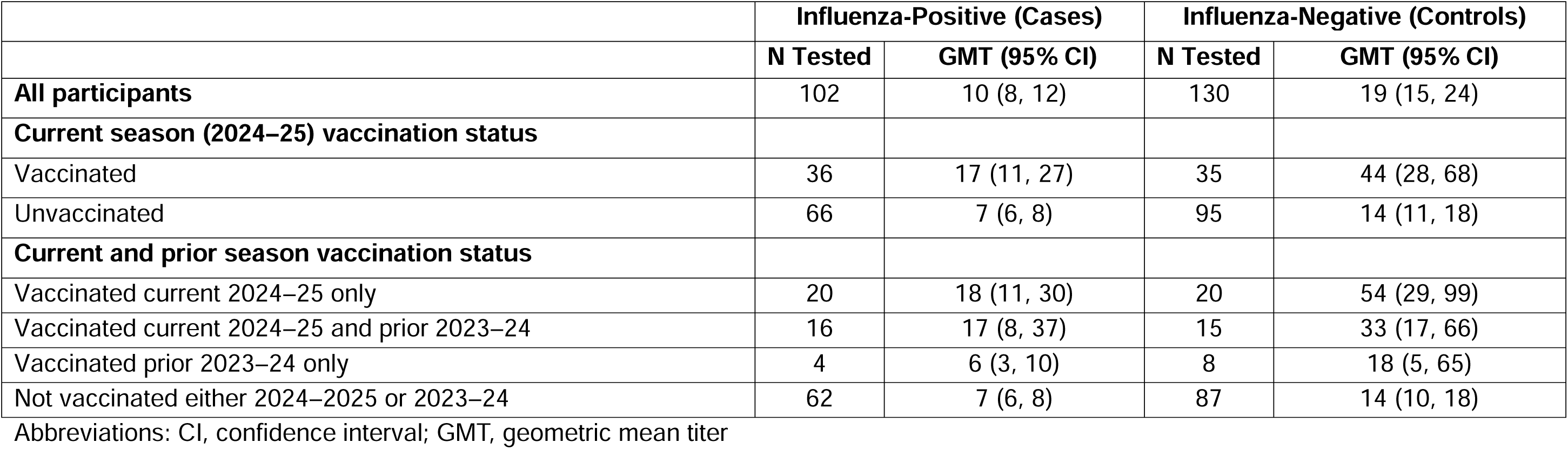
Microneutralization titers to A(H3N2) virus A/Wisconsin/154/2024 and 95% confidence intervals among laboratory-confirmed A(H3N2)-positive participants compared to test-negative participants aged ≥18 years by current and prior season influenza vaccination status.

**Supplemental Table 6.** Sensitivity analyses of influenza vaccine effectiveness against any influenza among participants of all ages.

|  | Influenza Positive (Cases) |  | Influenza Negative (Controls) |  | Vaccine Effectiveness<sup>a</sup> |  |
| --- | --- | --- | --- | --- | --- | --- |
|  | Total | No. Vaccinated (%) | Total | No. Vaccinated (%) | VE % | (95% CI) |
| **Primary analysis** | 575/2016 | 29 | 1687/4777 | 35 | 33 | (24 to 41) |
| **Varying exposure information source** |  |  |  |  |  |  |
| Defining vaccination status using self-reported information only | 450/1882 | 24 | 1239/4462 | 28 | 23 | (11 to 33) |
| Defining vaccination status using documented information only | 477/1964 | 24 | 1462/4707 | 31 | 37 | (28 to 45) |
| **Varying participant inclusion criteria** |  |  |  |  |  |  |
| Restricting to participants enrolled within 5 days of illness onset | 480/1674 | 29 | 1176/3471 | 34 | 32 | (21 to 41) |
| Restricting to participants enrolled with cough and fever | 437/1680 | 26 | 801/2519 | 32 | 39 | (28 to 47) |
| Include participants with known clinical results prior to enrollment | 759/2432 | 31 | 1922/5409 | 36 | 28 | (19 to 35) |
| Including SARS-CoV-2-positive participants | 576/2035 | 28 | 1828/5143 | 36 | 35 | (26 to 43) |
| Removing participants from Arizona | 550/1720 | 32 | 1592/4178 | 38 | 33 | (23 to 42) |
| Removing participants from Michigan | 529/1900 | 28 | 1508/4437 | 34 | 34 | (24 to 42) |
| Removing participants from Missouri | 558/1952 | 29 | 1554/4282 | 36 | 34 | (25 to 43) |
| Removing participants from Ohio | 511/1664 | 31 | 1526/4009 | 38 | 35 | (25 to 43) |
| Removing participants from Pennsylvania | 373/1433 | 26 | 1287/3846 | 33 | 35 | (24 to 44) |
| Removing participants from Texas | 438/1610 | 27 | 1235/3641 | 34 | 37 | (27 to 46) |
| Removing participants from Washington | 491/1817 | 27 | 1420/4269 | 33 | 35 | (25 to 43) |

<sup>a</sup> Models adjusted for study site, age, presence of $\geq 1$ underlying health condition, and month of illness onset. 95% confidence intervals that exclude 0% are considered statistically significant

## Supplemental Methods Microneutralization assay

For MN assays, serial 2-fold dilutions from an initial 1:10 dilution (lowest detectable MN titer) were mixed with influenza viruses (100 50% tissue culture infective doses [TCID50]). The mixtures were infected with 1.5 x 104 Madin-Darby canine kidney-SIAT1 (MDCK-SIAT1) cells per well, and viral infection was determined by ELISA. Neutralizing antibody titers were defined as the reciprocal of the highest dilution of serum samples that achieved at least 50% neutralization. Samples with titer results <10 were assigned a titer of 5. MN assay viruses included the predominantly detected A(H3N2) virus among the US Flu VE Network sequenced viruses: MDCK-SIAT1 grown A/Wisconsin/154/2024.

## Notes

### Author Declarations

The study protocol was reviewed by CDC, determined to be public health surveillance, and conducted consistent with applicable federal law and CDC policy (45 CFR 46.102(l)(2)).

## References

1. US Centers for Disease Control and Prevention. Flu Disease Burden: Past Seasons. January 14, 2025 (https://www.cdc.gov/flu-burden/php/data-vis/past-seasons.html).

2. US Centers for Disease Control and Prevention. Weekly US Influenza Surveillance Report: Key Updates for Week 19, ending May 10, 2025. (https://www.cdc.gov/fluview/surveillance/2025-week-19.html).

3. US Centers for Disease Control and Prevention. Influenza activity in the United States during the 2024-25 season and composition of the 2025-26 influenza vaccine. September 26, 2026 (https://www.cdc.gov/flu/whats-new/2025-2026-influenza-activity.html).

4. Grohskopf LA, Ferdinands JM, Blanton LH, Broder KR, Loehr J. Prevention and Control of Seasonal Influenza with Vaccines: Recommendations of the Advisory Committee on Immunization Practices - United States, 2024-25 Influenza Season. MMWR Recomm Rep 2024;73(5):1–25. (In eng). DOI: 10.15585/mmwr.rr7305a1.

5. US Centers for Disease Control and Prevention. CDC Seasonal Flu Vaccine Effectiveness Studies. May 30, 2025 (https://www.cdc.gov/flu-vaccines-work/php/effectiveness-studies/index.html).

6. US Centers for Disease Control and Prevention. Flu Burden Prevented by Vaccination: Past Seasons. June 26, 2025 (https://www.cdc.gov/flu-burden/php/data-vis-vac/past-burden-prevented-est.html).

7. Frutos AM, Cleary S, Reeves EL, et al. Interim Estimates of 2024-2025 Seasonal Influenza Vaccine Effectiveness - Four Vaccine Effectiveness Networks, United States, October 2024-February 2025. MMWR Morb Mortal Wkly Rep 2025;74(6):83–90. (In eng). DOI: 10.15585/mmwr.mm7406a2.

8. Chung JR, Price AM, Zimmerman RK, et al. Influenza vaccine effectiveness against medically attended outpatient illness, United States, 2023-24 season. medRxiv 2024 (In eng). DOI: 10.1101/2024.10.29.24316377.

9. US Centers for Disease Control and Prevention. People at increased risk for flu complications. September 11, 2024 (https://www.cdc.gov/flu/highrisk/index.htm).

10. Aksamentov I, Roemer C, Hodcroft EB, Neher RA. Nextclade: Clade Assignment, Mutation Calling and Quality Control for Viral Genomes. The Journal of Open Source Software 2021;6(67):3773. DOI: 10.21105/joss.03773.

11. World Health Organization. Manual for the laboratory diagnosis and virologic surveillance of influenza. WHO Press; 2011.

12. Gross FL, Bai Y, Jefferson S, Holiday C, Levine MZ. Measuring Influenza Neutralizing Antibody Responses to A(H3N2) Viruses in Human Sera by Microneutralization Assays Using MDCK-SIAT1 Cells. J Vis Exp 2017(129) (In eng). DOI: 10.3791/56448.

13. Jackson ML, Nelson JC. The test-negative design for estimating influenza vaccine effectiveness. Vaccine 2013;31(17):2165–8. (In eng). DOI: 10.1016/j.vaccine.2013.02.053.

14. Ramsay LC, Buchan SA, Stirling RG, et al. The impact of repeated vaccination on influenza vaccine effectiveness: a systematic review and meta-analysis. BMC Med 2019;17(1):9. (In eng). DOI: 10.1186/s12916-018-1239-8.

15. Kim SS, Flannery B, Foppa IM, et al. Effects of Prior Season Vaccination on Current Season Vaccine Effectiveness in the United States Flu Vaccine Effectiveness Network, 2012-2013 Through 2017-2018. Clin Infect Dis 2021;73(3):497–505. (In eng). DOI: 10.1093/cid/ciaa706.

16. Foppa IM, Ferdinands JM, Chung J, Flannery B, Fry AM. Vaccination history as a confounder of studies of influenza vaccine effectiveness. Vaccine X 2019;1:100008. (In eng). DOI: 10.1016/j.jvacx.2019.100008.

17. Doll MK, Pettigrew SM, Ma J, Verma A. Effects of Confounding Bias in Coronavirus Disease 2019 (COVID-19) and Influenza Vaccine Effectiveness Test-Negative Designs Due to Correlated Influenza and COVID-19 Vaccination Behaviors. Clin Infect Dis 2022;75(1):e564–e571. (In eng). DOI: 10.1093/cid/ciac234.

18. Okoli GN, Racovitan F, Abdulwahid T, Righolt CH, Mahmud SM. Variable seasonal influenza vaccine effectiveness across geographical regions, age groups and levels of vaccine antigenic similarity with circulating virus strains: A systematic review and meta-analysis of the evidence from test-negative design studies after the 2009/10 influenza pandemic. Vaccine 2021;39(8):1225–1240. (In eng). DOI: 10.1016/j.vaccine.2021.01.032.

19. Belongia EA, Simpson MD, King JP, et al. Variable influenza vaccine effectiveness by subtype: a systematic review and meta-analysis of test-negative design studies. Lancet Infect Dis 2016;16(8):942–51. (In eng). DOI: 10.1016/s1473-3099(16)00129-8.

20. Tenforde MW, Kondor RJG, Chung JR, et al. Effect of Antigenic Drift on Influenza Vaccine Effectiveness in the United States-2019-2020. Clin Infect Dis 2021;73(11):e4244–e4250. (In eng). DOI: 10.1093/cid/ciaa1884.

21. Hood N, Flannery B, Gaglani M, et al. Influenza Vaccine Effectiveness Among Children: 2011-2020. Pediatrics 2023;151(4) (In eng). DOI: 10.1542/peds.2022-059922.

22. Shinjoh M, Tamura K, Yamaguchi Y, et al. Influenza vaccination in Japanese children, 2024/25: Effectiveness of inactivated vaccine and limited use of newly introduced live-attenuated vaccine. Vaccine 2025;61:127429. (In eng). DOI: 10.1016/j.vaccine.2025.127429.

23. Grohskopf LA, Blanton LH, Ferdinands JM, Reed C, Dugan VG, Daskalakis DC. Prevention and Control of Seasonal Influenza with Vaccines: Recommendations of the Advisory Committee on Immunization Practices - United States, 2025-26 Influenza Season. MMWR Morb Mortal Wkly Rep 2025;74(32):500–507. (In eng). DOI: 10.15585/mmwr.mm7432a2.

24. DiazGranados CA, Dunning AJ, Kimmel M, et al. Efficacy of high-dose versus standard-dose influenza vaccine in older adults. N Engl J Med 2014;371(7):635–45. (In eng). DOI: 10.1056/NEJMoa1315727.

25. Nicolay U, Heijnen E, Nacci P, Patriarca PA, Leav B. Immunogenicity of aIIV3, MF59-adjuvanted seasonal trivalent influenza vaccine, in older adults ≥65 years of age: Meta-analysis of cumulative clinical experience. Int J Infect Dis 2019;85s:S1–s9. (In eng). DOI: 10.1016/j.ijid.2019.03.026.

26. Ferdinands JM, Blanton LH, Alyanak E, et al. Protection against influenza hospitalizations from enhanced influenza vaccines among older adults: A systematic review and network meta-analysis. J Am Geriatr Soc 2024;72(12):3875–3889. (In eng). DOI: 10.1111/jgs.19176.

27. Lewis NM, Harker EJ, Cleary S, et al. Vaccine Effectiveness Against Influenza A(H1N1), A(H3N2), and B-Associated Hospitalizations, United States, 1 September 2023 to 31 May 2024. The Journal of Infectious Diseases 2025;232(4):e626–e636. DOI: 10.1093/infdis/jiaf185.

28. Belongia EA, Skowronski DM, McLean HQ, Chambers C, Sundaram ME, De Serres G. Repeated annual influenza vaccination and vaccine effectiveness: review of evidence. Expert Rev Vaccines 2017;16(7):1–14. (In eng). DOI: 10.1080/14760584.2017.1334554.

29. Jones-Gray E, Robinson EJ, Kucharski AJ, Fox A, Sullivan SG. Does repeated influenza vaccination attenuate effectiveness? A systematic review and meta-analysis. Lancet Respir Med 2023;11(1):27–44. (In eng). DOI: 10.1016/s2213-2600(22)00266-1.

30. Bartoszko JJ, McNamara IF, Aras OAZ, et al. Does consecutive influenza vaccination reduce protection against influenza: A systematic review and meta-analysis. Vaccine 2018;36(24):3434–3444. (In eng). DOI: 10.1016/j.vaccine.2018.04.049.

31. Leis AM, Wagner A, Flannery B, Chung JR, Monto AS, Martin ET. Evaluation of test-negative design estimates of influenza vaccine effectiveness in the context of multiple, co-circulating, vaccine preventable respiratory viruses. Vaccine 2024;42(26):126493. (In eng). DOI: 10.1016/j.vaccine.2024.126493.

32. Lewis NM, Harker EJ, Leis A, et al. Assessment and mitigation of bias in influenza and COVID-19 vaccine effectiveness analyses - IVY Network, September 1, 2022-March 30, 2023. Vaccine 2025;43(Pt 2):126492. (In eng). DOI: 10.1016/j.vaccine.2024.126492.

33. Goldsmith JJ, Tavlian S, Vu C, Regan AK, Campbell PT, Sullivan SG. Comparison of 2 Doses vs 1 Dose in the First Season Children Are Vaccinated Against Influenza: A Systematic Review and Meta-Analysis. JAMA Netw Open 2025;8(10):e2535250. (In eng). DOI: 10.1001/jamanetworkopen.2025.35250.

34. Chung JR, Rolfes MA, Flannery B, et al. Effects of Influenza Vaccination in the United States During the 2018-2019 Influenza Season. Clin Infect Dis 2020;71(8):e368–e376. (In eng). DOI: 10.1093/cid/ciz1244.

35. Rolfes MA, Flannery B, Chung JR, et al. Effects of Influenza Vaccination in the United States During the 2017-2018 Influenza Season. Clin Infect Dis 2019;69(11):1845–1853. (In eng). DOI: 10.1093/cid/ciz075.

